# Understanding the psychological journey of patients with head and neck cancer in different periods: a qualitative study protocol

**DOI:** 10.1101/2021.04.27.21256227

**Authors:** Yin Li, Cheng Ping, Li Yijian, Yang Chunlian, Jiang Ren, Yang Jing, Liang Haixin, Jiang Qinghua, Lyu Jianxia

## Abstract

**Introduction:** There are more than 900,000 new cases of head and neck tumors in the world every year, which is the sixth most common tumor in the world. During the treatment of head and neck cancer, patients often face various symptoms and psychological problems. In this qualitative study, patients with head and neck cancer were invited to share their psychological journey during diagnosis, treatment, and recovery to (1) understand their psychological status and feelings during different treatment stages, (2) their needs and concerns during different treatment stages, and (3) difficulties and problems in self-management.

**Methods and analysis:** We adopt qualitative study to understand these patient’ psychological journey with head and neck cancer, especially when they are at different times in the diagnosis, treatment and completion of head and neck tumors. Thematic analysis will be carried out following the six-phase iterative process suggested by Braun and Clarke: (1) familiarising oneself with data, (2) generating initial codes, (3) searching for themes, (4) reviewing themes,(5) defining and naming themes and (6) providing the report, which will be used to analyse the qualitative data.

**Ethics and dissemination:** This study has ethical approval from the Sichuan Cancer Hospital Ethics Committee. We will disseminate the findings through national and international conferences and international peer-reviewed journals.

**Strengths and limitations:** In this study, qualitative research was used to better understand the psychological process and feelings of patients with head and neck tumor at different stages of treatment.

The subjects were interviewed in the same tumor hospital. Patients in different grade hospitals in different regions may have different psychological processes and experiences.

## INTRODUCTION

Head and Neck Cancer (HNC) is a tumor that originates in various parts of the head and neck area, including the oral cavity, pharynx, larynx, nasal cavity, and paranasal sinuses and glands. It can involve the eyes, ears, nose, throat, and thyroid^1^. According to statistics, there are more than 900,000 new cases of head and neck cancer every year^2^, which is the sixth most common type of cancer in the world today. China is a big country of head and neck cancer, with an annual incidence of 15.34/100,000^3^, accounting for about 10% of systemic malignant cancers. With the advancement of medical standards, the survival rate of head and neck tumors continues to extend. The 5-year overall survival rate of head and neck tumors after standardized treatment is about 80% (China 92 staging)^4^, but the anatomical structure of head and neck tumors is relatively complex, involving important tissues, organs and nerves in the head and neck, causing clinical symptoms. At present, patients with head and neck cancer mostly adopt a comprehensive treatment strategy of radiotherapy, chemical and molecular targeting^5^. Intensity-modulated radiotherapy (IMRT) and a diverse combination of chemotherapy, concurrent radiotherapy and chemotherapy have improved the control rate and overall survival rate of head and neck cancers^6^. Radiation reactions and radiation damage caused by treatment, side effects of chemotherapy and gastrointestinal reactions, effectiveness and safety of targeted therapy are the main factors affecting patients’ willingness to treat and quality of life.

As an important part of the human body, the head and neck bear hearing, smell and taste, control people’s chewing, swallowing, breathing, language expression and other physiological functions, and have a great impact on people’s appearance image^7^. Compared with other cancers, patients with head and neck cancer suffer greater emotional trauma^8^. Due to the complexity and particularity of head and neck anatomy parts, patients in addition to bring the symptoms of the disease itself, such as nasal congestion, chewing difficulty swallowing or breathing difficulties, will also suffer from surgery, chemotherapy and radiotherapy treatments such as all kinds of adverse reactions, such as dry mouth, oral mucositis, radioactive dermatitis, difficulty swallowing, even appearance defects, etc. According to statistics^9^, among the adverse reactions of radiotherapy in the head and neck, oral mucosal reaction is up to 97%, dry mouth is 60% ∼ 90%, sensorineural deafness is 40% ∼ 60%, grade 3 dysphagia is 15% ∼ 30%, and osteonecrosis of the jaw is 5% ∼ 15%. These symptoms and side effects seriously affect the physical and mental health of patients. Patients may also suffer from a decrease in self-image level, which is often accompanied by unwillingness to communicate with the outside world, stigma, depression^10-12^, and ultimately lead to a decline in the quality of life of patients^13^. Assessing whether a patient is experiencing physical or psychological symptoms and then providing appropriate help as needed is critical to improving their quality of life.

Qualitative research is an activity that takes the researcher himself as the research tool, adopts a variety of data collection methods to comprehensively explore social phenomena in natural situations, analyzes data by induction, and gets explanatory understanding of the behavior and meaning construction of research objects through interaction with them^14^. Nursing is an applied discipline with strong humanism, and the practice field of nursing profession always revolve around people, while qualitative research focuses on people’s real feelings and experience, which is interlinked with the characteristics of nursing profession. Therefore, this study will adopt the qualitative research method to understand the psychological process and feelings of patients with head and neck tumor in different stages of diagnosis, diagnosis and treatment and rehabilitation, to provide reference for clinical medical staff to provide patients with symptomatic support nursing.

## METHODS

### Objectives

The objectives of this study are to:

1. To explore the experience of patients with head and neck cancer in the initial diagnosis, treatment and rehabilitation of the disease.
2. To explore the main content and focus of attention of patients with head and neck cancer in different treatment stages of supportive care needs.
3. What are the difficulties and problems of self-management in patients with head and neck cancer.

### Study design and setting

Qualitative study and semi-structured in-depth interview method were used to explore the psychological process, feelings and care needs of patients with head and neck cancer at admission, during treatment, at discharge, and at the first review.

Objective sampling was used to consider the severity of disease, cognitive function, emotional status and symptoms of patients. If the patient has slurred speech and speech difficulties due to oral pain or brain tumor, the main family caregivers are interviewed.

The inclusion criteria were as follows: (1) Age 18 or above; (2) Any nationality or sex; 3) Patients with head and neck tumors requiring radiotherapy and chemotherapy; (4) Radiotherapy range includes primary lesion and neck lymphatic drainage area; (5) Clear mind, no communication barriers, normal cognitive function, able to answer questions; (6) Signing the informed consent.

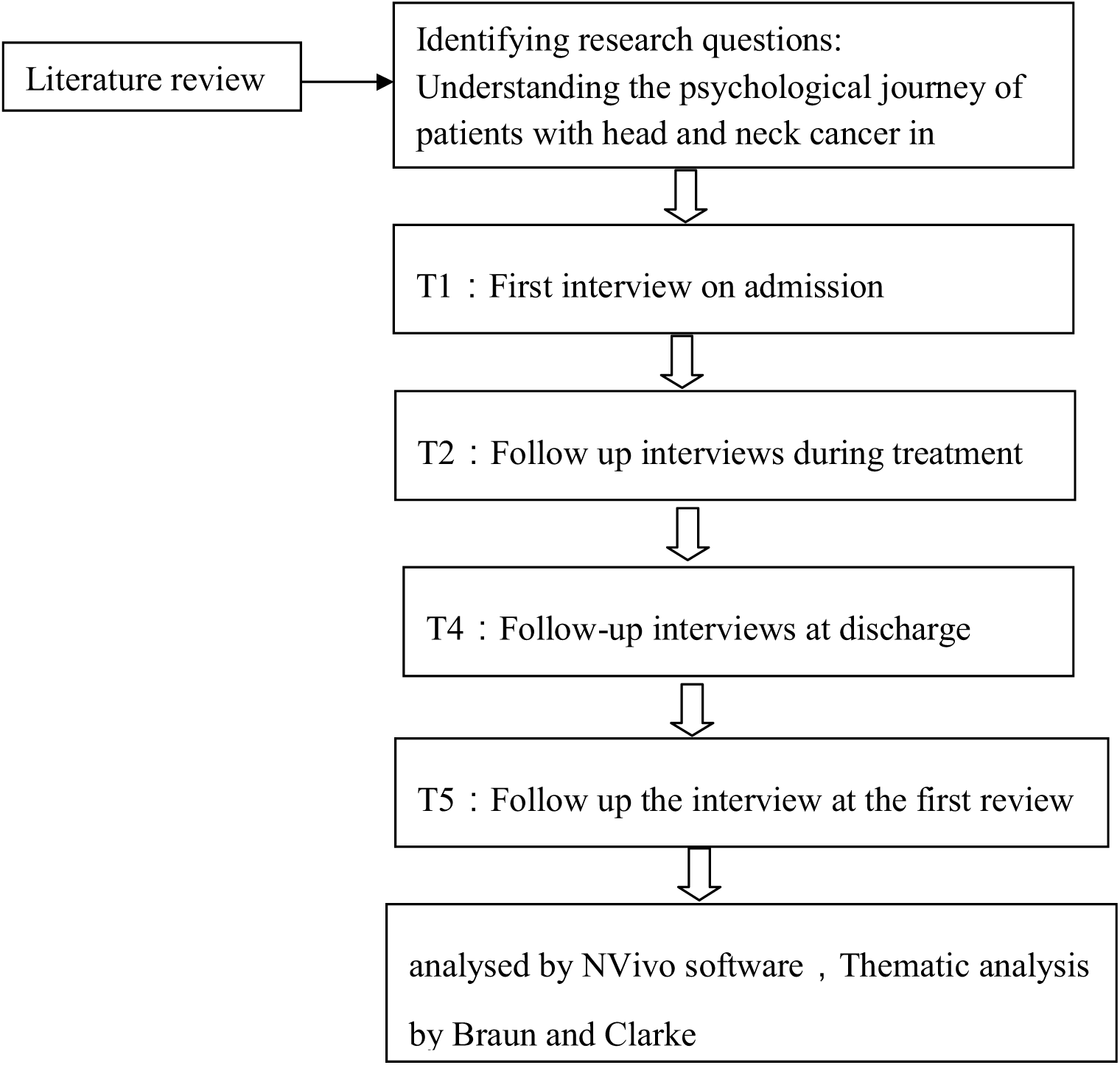

### Data collection

After the patient was admitted to the hospital, a good nurse-patient relationship was established with the patient and family members, and the interview began after obtaining informed consent. Around the beginning of the interview outline, the interview was recorded by a combination of recording and notes, and non-verbal behaviors such as the posture, tone, and facial expressions of patients and family members were recorded. Each interview takes about 40-60 minutes. Before the end of the interview, ask the patient and family members if there is any content that needs to be added, and thank you. the interview outline we worked out is as follows: 1). Can you talk a little bit about how you’ve felt since you discovered your illness? / How have you felt since the beginning of the treatment (since the end of the treatment)? 2) What problems/difficulties did you encounter during this process? 3) How have these problems/difficulties affected you and your family? 4) How did you solve these problems/difficulties?Do you feel that you and your family have enough ability to solve these problems? 5) What are your needs (information, functional exercises, support, etc.) 6) What kind of support and help do you expect?From where?By whom?How to provide.

### Data analysis

The audio recording of the semi-structured interviews will be transcribed in full, stored and analysed using NVivo 12pro software. Thematic analysis will be carried out following the six-phase iterative process suggested by Braun and Clarke^15^: (1) familiarising oneself with data, (2) generating initial codes, (3) searching for themes, (4) reviewing themes,(5) defining and naming themes and (6) providing the report, which will be used to analyse the qualitative data ^16-18^.Our analysis will adopt an inductive (‘bottom up’) approach (ie, without integrating the themes within any pre-existing coding schemes or preconceptions of the researchers) to identify patterned meanings in the data set within an constructivism framework (to report the experiences, meanings and reality of the participants).

## Data Availability

The datasets generated and/or analysed in the current study are not publicly available due to individual privacy but are available from the corresponding author on reasonable request.

## Ethical considerations

All patients signed informed consent forms. This study was approved by the Sichuan Cancer Hospital Ethics Committee (SCCCIIEC-02-02020-065). Three specific ethical issues were highlighted, including (1) confidentiality and anonymity, (2) power dynamics and (3) researching potentially sensitive topics

## Dissemination

The researchers will prepare manuscripts and publish the results from this study in relevant peer-reviewed journals and present findings via poster presentations and oral presentations at appropriate academic and non-scientific conferences.

## Acknowledgements

The authors would like to thank every patient and family member who participated in the study.

## Author’s information

Yin Li is a master of nursing and a head nurse in the Head and Neck Radiotherapy Department of Sichuan Cancer Hospital. And she also has 23 years of clinical oncology nursing and management experience, focusing on the symptoms and clinical problems of patients undergoing head and neck radiotherapy.

Lyu Jianxia is a Master of Nursing and a clinical nurse in the Head and Neck Radiotherapy Department of Sichuan Cancer Hospital, mainly engaged in clinical and psychological nursing of cancer patients.

## Authors’ contributions

JXL and LY contributed to the design, interview the participants and manuscript writing. JY, HXL and QHJ devised and piloted the interview content and procedures. PC, YJL, CLY and RJ recruited the participants. All authors read and approved the final manuscript.

## Funding

The study was supported by a financial grant from the Sichuan Provincial Health Research Project of the Sichuan Provincial Health Committee, NO. 19PJ142, Science and technology department of Sichuan Province 2020 research project, NO. 2020JDRC0121 and NO. 2020YFS0411. Funders had no direct role in developing this piece of work and all opinions expressed are of the authors as individuals and not on behalf of their institutions.

## Competing interests

None declared.

## Patient consent for publication

Not required.

## Provenance and peer review

Not commissioned; externally peer reviewed.

## Notes

### Competing Interest Statement

The authors have declared no competing interest.

### Author Declarations

This study was approved by the Sichuan Cancer Hospital Ethics Committee (SCCCIIEC-02-02020-065).

## References

1 Arboleda LPA, D. MendonÇa RMH, Lopez EEM, et al. Global frequency and distribution of head and neck cancer in pediatrics:a systematic review[J]. Crit Rev Oncol Hematol, 2020,148:102892.

2 Garg S,Yoo J, Winquist E. Nutritional support for head and neck cancer patients receiving radiotherapy: a systematic review[J]. Support Care Cancer, 2010,18(6):667–677.

3 Bray F, Ferlay J, Soerjomataram I, et al. Global cancer statistics 2018: GLOBOCAN estimates of incidence and mortality worldwide for 36 cancers in 185 countries[J]. CA Cancer J Clin, 2018,68(6):394–424.

4 Sun Y. Clinical Oncology.Beijing: China Medical Electronic Audio & Video Press,2017:357–373.

5 Shellenberger T D, Weber R S. Multidisciplinary team planning for patients with head and neck cancer[J]. Oral Maxillofac Surg Clin North Am, 2018,30(4):435–444.

6 Brunner M, Gore S M, Read R L, et al. Head and neck multidiplinary team meetings: effect on patient management[J]. Head and Neck, 2015,37(7):1046–1050.

7 Wang Mengyao, Wang Binquan, Wang Lei, et al. Summary of nursing guidelines for head and neck cancer survivors, CHINESE NURSING RESEARCH January, 2021,35(1):63–66.

8 Björklund M, Sarvimäki A, Berg A, et al. Living with head and neck cancer: a profile of captivity[J]. Journal of Nursing and Healthcare of Chronic Illness, 2010,2(1):22.

9 Chinese Society of Radiation Oncology of Chinese Medical Association. Expert consensus on prevention and control strategy of radiotherapy-induced oral mucositis (2019) [J]. J Oral Oncol, 2012,28(9):641–647.

10 Chen S C, Yu P J, Hong M Y, et al. Communication dysfunction, body image, and symptom severity in postoperative head and neck cancer patients: factors associated with the amount of speaking after treatment[J]. Support Care Cancer, 2015,23(8):2375–2382.

11 Kam D, Salib A, Gorgy G, et al. Incidence of Suicide in Patients With Head and Neck Cancer[J]. JAMA Otolaryngol Head Neck Surg, 2015,141(12):1075–1081.

12 Kissane D W, Patel S G, Baser R E, et al. Preliminary evaluation of the reliability and validity of the Shame and Stigma Scale in head and neck cancer[J]. Head Neck, 2013,35(2):172–183.

13 CAO Jiayan, CHEN Changlian, WANG Lingling, et al. Analysis on body image disorder in patients with head and neck cancer and its influencing factors[J]. Chinese Nursing Management Vol.19, No.09, September 15, 2019:1317-1321.

14 LI Zheng, LIU Yu. Nursing research methods [M]. Beijing: People’s Medical Publishing House, 2018.

15 Braun V, Clarke V. Using thematic analysis in psychology. Qual Res Psychol 2006;3:77–101.

16 Braun V, Clarke V. Reflecting on reflexive thematic analysis. Qual Res Sport Exerc Health 2019;11:589–97.

17 Clarke V, Braun V. Thematic analysis. In: Lyons E, Coyle A, eds. Analysing qualitative data in psychology. London, UK: SAGE, 2016

18 Analysing qualitative data in psychology. London, UK: SAGE, 2016. Braun V, Clarke V, Hayfield N, et al. Thematic analysis. In: Liamputtong P, ed. Handbook of research methods in health social sciences. Singapore: pnSpringer Singapore, 2019: 843–60.

